# Impact of antenatal care quality on neonatal mortality in Ethiopia: Evidence from DHS (2000 to 2019)

**DOI:** 10.1101/2025.06.01.25328760

**Authors:** Fikreselassie Getachew, Ashenif Tadele, Mulugeta Gajea, Girum Taye, Hiwot Achamyeleh, Asrat Arja, Shegaw Mulu, Getachew Tollera, Dessalegn Y Melesse

**Author notes:** **Co-authors:** Ashenif Tadele^1^, Mulugeta Gajea^3^, Girum Taye^1^, Hiwot Achamyeleh^1^, Asrat Arja^1^, Shegaw Mulu^4^, Aderajew Mekonnen^1^, Getachew Tollera^1^, Dessalegn Y Melesse^5^.

## Abstract

**Background:** Globally, 2.3 million children died, with an average of 6,300 neonatal deaths every day. Sub-Saharan Africa (SSA) has the world’s highest infant death rate and has made the slowest progress in reducing neonatal mortality. Studies on the impact of antenatal care quality on neonatal mortality in Ethiopia have been limited.

**Method:** This study analyzed five years of Ethiopian demographic and health survey data, examining 51,730 live births from the five years prior to the survey. It assessed the quality of antenatal care through six key components. Utilizing multivariate logistic regression, the study explored the relationship between antenatal care quality scores, ANC visits, service components, and neonatal mortality. Factors with p-values below 0.05 were deemed statistically significant.

**Results:** The quality of prenatal care was 2.69 in 2016, ranging from 2.25 in the Somali region to 6.35 in Addis Ababa, 5.36 in higher education, 4.12 among the wealthiest individuals, and 4.7 in urban subpopulations. Neonatal mortality decreased by 16.3% for each unit increase in prenatal care quality. The reduction in neonatal mortality was associated with AN4+ (AOR=0.46, 95%CI 0.33, 0.64), good quality of antenatal care (AOR=0.56, 95% CI 0.42, 0.74), blood pressure measurements (AOR= 0.42, 95% CI 0.23, 0.75), and counseling about pregnancy complications (AOR= 0.58, 95% CI 0.35, 0.95).

**Conclusion:** The Quality of antenatal care varies across regions, and is more advantageous in the subpopulations. Reducing neonatal mortality is associated with pregnant women receiving quality ANC, attending at least four ANC visits, having blood pressure measurements, and receiving advice on pregnancy-related difficulties.

## Background

Neonatal mortality is defined as a newborn’s death during the first four weeks of life (the neonatal period), and it is measured in neonatal deaths per 1000 live births (1). Neonatal mortality remains a significant global issue, particularly prevalent in Low and Middle-Income Countries (LMICs) such as Ethiopia. Neonatal mortality remains a major global issue, especially in low- and middle-income countries like Ethiopia. In 2022, an estimated 2.3 million neonatal deaths occurred worldwide, making up 47% of all under-five child deaths (1,2).

Saharan Africa (SSA) showed the highest neonatal mortality and the lowest progress has shown in reducing neonatal mortality (3). According to the Ethiopian Demographic and Health Survey (MINI-EDHS 2019), Neonatal Mortality was reported to be 33 deaths per 1000 live births in Ethiopia (4).

Evidence indicated that nearly 75% of neonatal deaths in developing countries could be prevented with simple and existing low-cost tools (5). One of the main approaches recommended to lower the risk of neonatal mortality is ANC follow-up, which is applied in every population regardless of socioeconomic status (6,7). Antenatal care (ANC) follow-up increases the chances of a newborn’s survival and health by directly reducing neonatal deaths and indirectly providing mothers with access to health services. For instance, a study in Zimbabwe found that a one-unit increase in the quality of prenatal care reduced the risk of neonatal mortality by nearly 42% (8). It indicated that giving attention to quality is crucial rather than focusing solely on access to ANC care utilization.

Ethiopia is now one of the countries experiencing the worst neonatal health outcomes in sub-Saharan Africa (SSA) (9), underscoring the critical need to understand the link between prenatal care quality and child health outcomes. Ending preventable neonatal deaths is one of the United Nations’ 2015 Sustainable Development Goals (SDGs) that aim to reduce neonatal mortality to 12 deaths per 1,000 live births by 2030 (10).

Antenatal care follow-up is one of the most important interventions for reducing the risk of neonatal mortality in any community, regardless of socioeconomic status (11). It directly improves the survival and health of children by lowering stillbirths and neonatal mortality, and indirectly by giving an entrance point for health interactions with the mother at a critical moment in the care continuum (12).

While several studies globally have examined the relationship between ANC follow-up and neonatal mortality, research focusing on the impact of quality ANC on neonatal mortality remains rare. For instance, a systematic review and meta-analysis study in Debre Markos, Ethiopia, revealed that ANC follow-up could reduce neonatal mortality by 34%. However, the study did not evaluate the impact of ANC quality (5).

Therefore, this study aimed to determine the effect of ANC Quality on neonatal mortality by using five EDHS surveys on neonatal mortality in Ethiopia.

## Methods

### Data sources

We utilized secondary data from five rounds of the Ethiopia Demographic and Health Survey (EDHS), conducted in 2000, 2005, 2011, 2016, and 2019. These data were accessed from the official page of the DHS program database (www.measuredhs.com) after receiving authorization via an online request, in which the objectives of our study were clearly outlined. The DHS survey is a nationally representative household survey that offers data from a wide variety of populations, health, and nutrition tracking, and effect assessment measures with face-to-face interviews of women aged 15–49 and their children Detailed survey techniques and methods of sampling are used to collect data have been recorded elsewhere (13).

A stratified two-stage cluster sampling scheme was used. The first stage comprised a random sampling of enumeration areas, while the second stage comprised a random sampling of households (13). The research utilized the birth data file from the EDHS dataset, from which both dependent and independent variables were extracted

### Population and eligibility criteria

The source population for this study included all pregnant women aged 15–49 who had the most recently given live birth in Ethiopia. The study population comprised all pregnant women aged 15–49 in Ethiopia who were selected as part of the sample. Pregnant women who did not receive all nine components of ANC services for their most recent birth were excluded from the study. Accordingly, a total of 48,678 pregnant women who received ANC at least once during their most recent pregnancy and who had all the components of ANC services were included in this study.

### Measurements of neonatal mortality and antenatal care quality

The EDHS gathers detailed birth histories from every interviewed woman in the reproductive age group of 15–49 years. The outcome variable for this study was neonatal mortality, which is binary-coded: 1 if the child died before reaching 28 days (neonatal period) and 0 otherwise. The explanatory variables included the quality of antenatal care (ANCq), the number of ANC visits, antenatal care, skilled birth attendance, place of birth, ANC provider, and antenatal components.

We define the quality of ANC as the receipt of all essential components of ANC services, including blood pressure checks, urine sample tests, blood sample tests, tetanus injections, weight measurements, height measurements, drugs for intestinal parasites, tetanus injections before birth, and counseling about pregnancy complications during pregnancy.

The construction of ANCq was guided by the WHO antenatal care guidelines (14). We created an overall measure to ascertain if the women received all six essential components of ANC. These services included (1) blood pressure checks, (2) urine sample tests, (3) blood sample tests, (4) tetanus injection, (5) weight measurements, (6) height measurements, (7) drugs for intestinal parasites, (8) tetanus injections before birth and (9) counsel about a pregnancy complication. Each question has a binary response (1 = Yes, and 0 = No). We coded each response as one if a particular service was received and zero otherwise. Following Deb and Sosa-Rubi (15). We created an index to measure the quality of antenatal care by adding up all the “yes” responses for each woman. ANCq is categorized as good or poor. The ANC score is poor when ANC. Score is 0-4, else (5-9)-good.

### Data processing and analysis

Data processing and analysis were conducted using STATA 16 software. Data were weighted using sampling weight, primary sampling unit, and strata before any statistical analysis to account for the unequal probability sampling in different strata and to ensure the representativeness of the findings. To standardize expected clinical actions, the analysis focused on women attending their first ANC visit. Cases with incomplete data were excluded. Cross-tabulations and summary statistics were used to describe the study population.

EDHS sample weights were used throughout the analysis. We used logistic regression to analyze the relationship between our proposed score for quality coverage, complete for ANC, and each ANC recommended clinical actions with neonatal mortality, estimating an odds ratio (OR). Bivariate and multivariate logistic regression were done to assess the association of each independent variable with the outcome, and crude and adjusted odds ratios, 95% confidence interval (CI), and p-values were presented. Independent variables found significant at the bivariable level with p-values less than 0.25 were added to the multivariable logistic regression model. Adjusted odds ratios (AOR), 95% Confidence Intervals (CI), and p-values were calculated with a statistical significance level set at p-value < 0.05.

Two models were fitted. The first was the multivariable model adjusted for maternal characteristics to verify the association of quality of antenatal care, antenatal visit count, place of delivery, and delivery assisted by skilled personnel with neonatal mortality. The second model was adjusted for ANC components to assess the association of ANC components with neonatal mortality.

### Ethical approval

Ethical approval was obtained from the Ethiopian Public Health Institute Institutional Review Board (IRB). The datasets were requested online, and permission to use the data was obtained from DHS.

## Results

### Background characteristics of study participants

This study included a total of 48,678 women who gave birth in the preceding five years. The mean age of the women was found to be 29.3 ± 6.84 years with a 95% CI: (29.28–29.40). The majority of women (71%) were within the age group of 20 to 34 years. Most of the women resided in rural areas and the Oromia region (88%) and (41.4%), respectively. Around 92% of them were married, and seven to ten women had no education (**Table 1**).

**Table 1:**
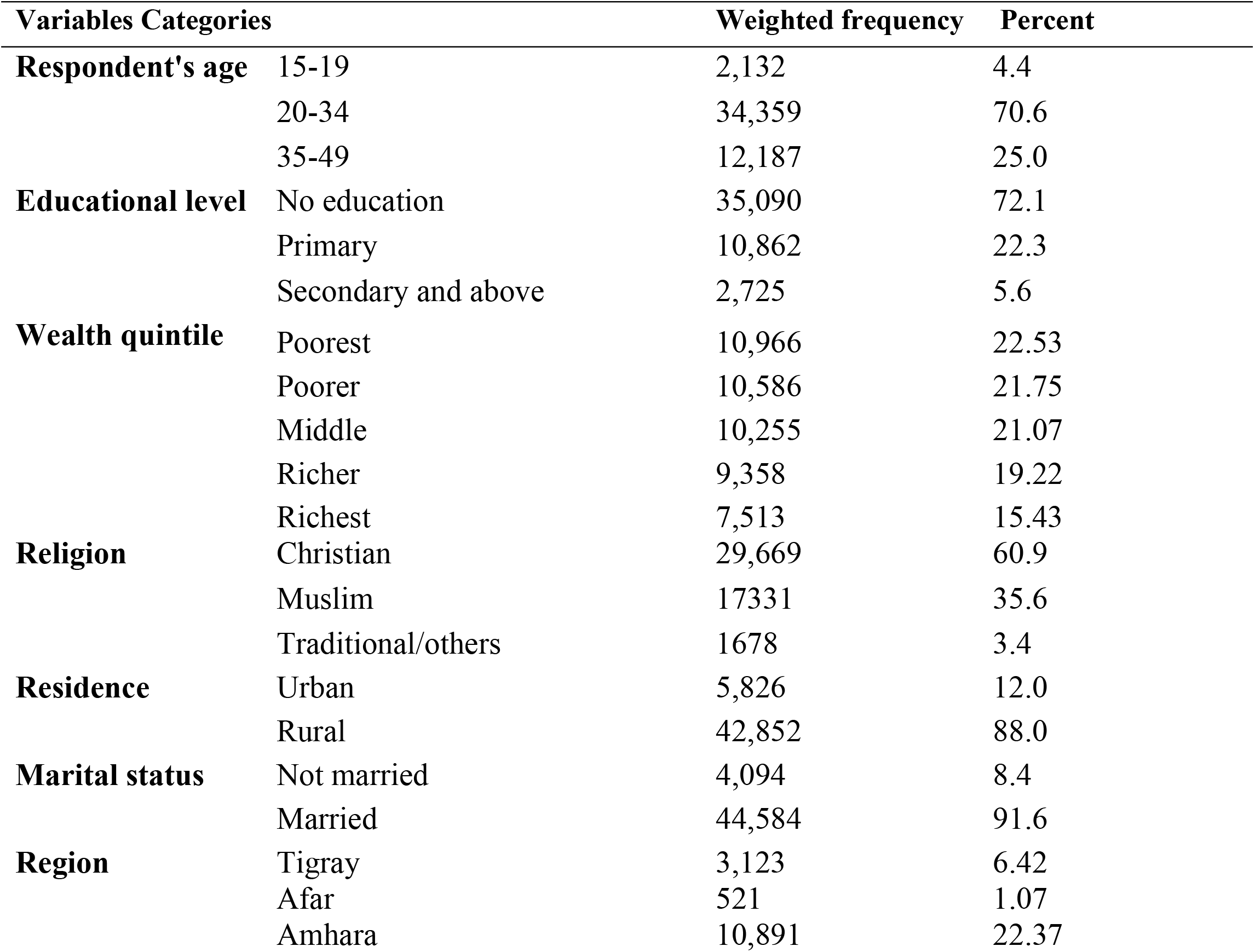
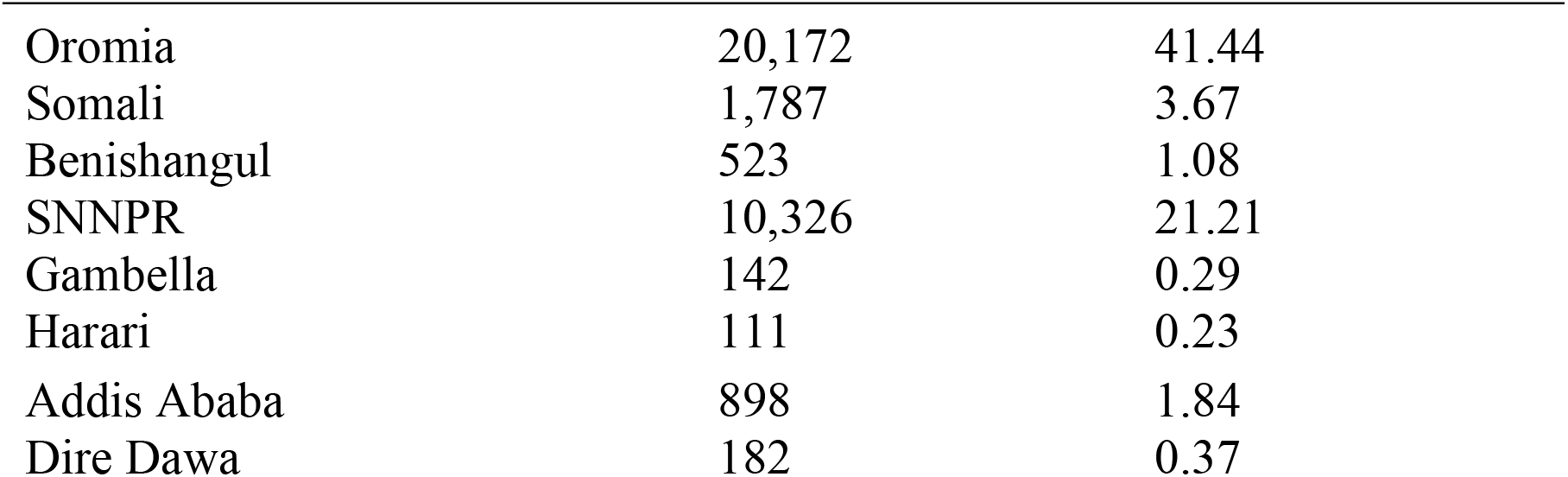
Background characteristics of study participants, Ethiopian DHS from 2000-2019.

| Variables Categories |  | Weighted frequency | Percent |
| --- | --- | --- | --- |
| Respondent's age | 15-19 | 2,132 | 4.4 |
|  | 20-34 | 34,359 | 70.6 |
|  | 35-49 | 12,187 | 25.0 |
| Educational level | No education | 35,090 | 72.1 |
|  | Primary | 10,862 | 22.3 |
|  | Secondary and above | 2,725 | 5.6 |
| Wealth quintile | Poorest | 10,966 | 22.53 |
|  | Poorer | 10,586 | 21.75 |
|  | Middle | 10,255 | 21.07 |
|  | Richer | 9,358 | 19.22 |
|  | Richest | 7,513 | 15.43 |
| Religion | Christian | 29,669 | 60.9 |
|  | Muslim | 17331 | 35.6 |
|  | Traditional/others | 1678 | 3.4 |
| Residence | Urban | 5,826 | 12.0 |
|  | Rural | 42,852 | 88.0 |
| Marital status | Not married | 4,094 | 8.4 |
|  | Married | 44,584 | 91.6 |
| Region | Tigray | 3,123 | 6.42 |
|  | Afar | 521 | 1.07 |
|  | Amhara | 10,891 | 22.37 |

|  |  |  |
| --- | --- | --- |
| Oromia | 20,172 | 41.44 |
| Somali | 1,787 | 3.67 |
| Benishangul | 523 | 1.08 |
| SNNPR | 10,326 | 21.21 |
| Gambella | 142 | 0.29 |
| Harari | 111 | 0.23 |
| Addis Ababa | 898 | 1.84 |
| Dire Dawa | 182 | 0.37 |

### Trends of ANC visits components

From 2000 to 2019, blood pressure measurements during ANC visits increased from 69% to 88 %, blood samples taken increased from 24.8% to 78.9%, and ANC visits by skilled providers rose from 3.2% to 32.0%. The pooled estimate from the 2000 to 2019 DHS survey indicated that the most frequently received components during pregnancy were blood pressure measurements (74.2%), blood samples (56.6%), and urine samples (50.4%). In contrast, the least common antenatal service item received was skilled delivery ANC (10.6%) (**Table 2**).

**Table 2:** Components of ANC service utilization from 2000 to 2019 DHS surveys.

| ANC components | 2000 | 2005 | 2011 | 2016 | 2019 | Pooled |
| --- | --- | --- | --- | --- | --- | --- |
| ANC by Skilled birth attendant | 3.2 | 3.5 | 7.1 | 19.2 | 31.9 | 10.6 |
| Blood pressure taken | 69.3 | 61.9 | 71.6 | 75.2 | 88.1 | 74.2 |
| Blood sample taken | 24.8 | 26.2 | 54 | 72.5 | 78.9 | 56.6 |
| Urine sample taken | 21.4 | 26.6 | 41.3 | 66.1 | 73.9 | 50.4 |

### ANC quality score: Pooled estimates and trends

Due to the absence of some ANC components in the MINI EDHS 2019 survey, the pooled ANCq score was calculated using data from the four EDHS surveys conducted between 2000 and 2016. The national pooled ANCq was 2.69, ranging from 2.25 in the Somali region to 6.35 in Addis Ababa. Women from advantageous subpopulations, such as those with higher education, the wealthiest, and urban residents, had better ANCq scores of 5.36, 4.12, and 4.7, respectively (**Figure 1**).

**Figure 1:**
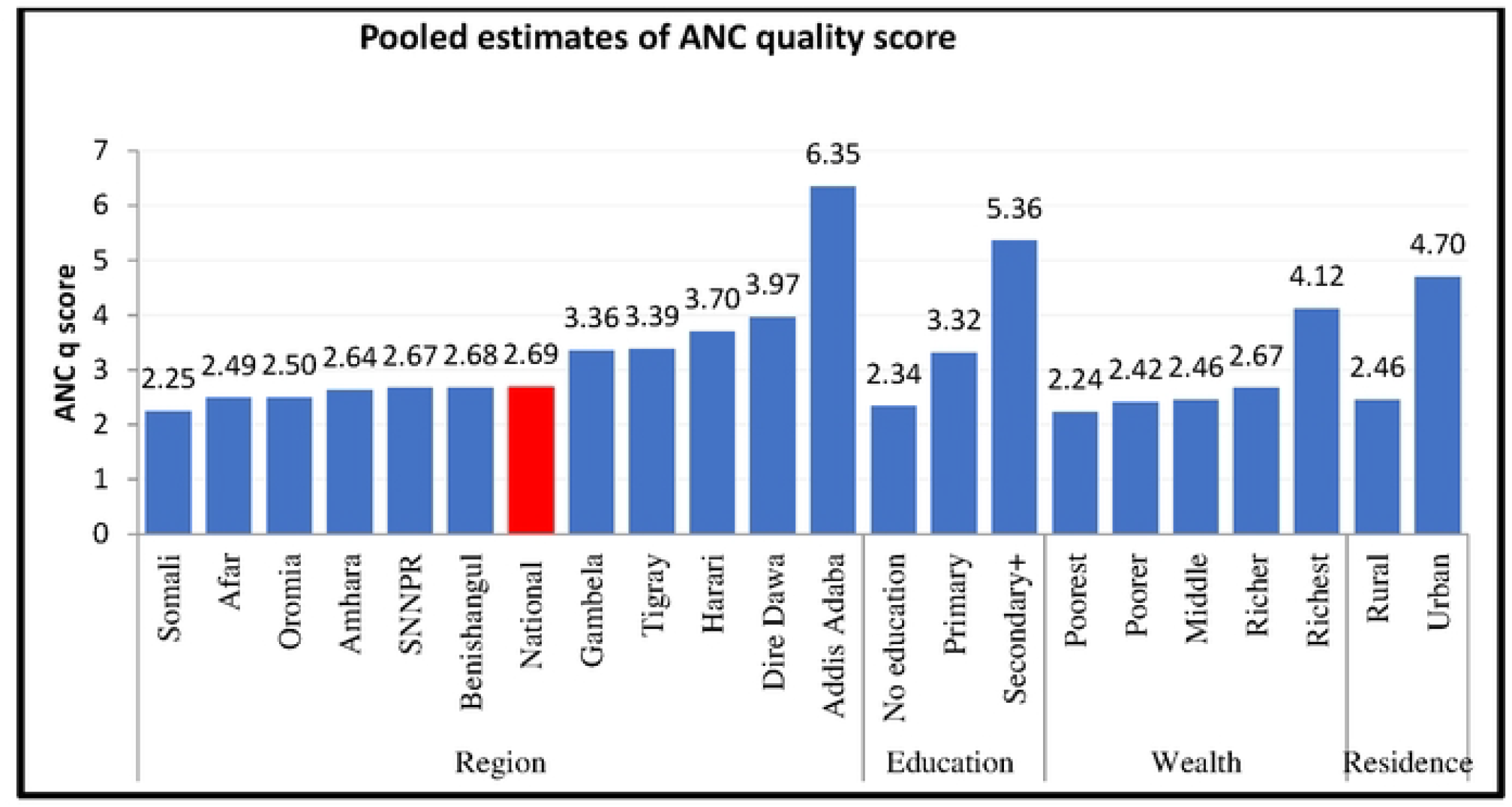
Pooled ANCq score distribution by background characteristics.

Nationally, the ANCq score has changed from 2.48 in 2000 to 2.69 in 2016 with an average annualized rate of Increment change (AARI) of 4.74%. Both Addis Ababa and Dire Dawa cities showed a 1.55% and 1.83% average annual rate of increment (AARI) from the year 2000 to 2016 DHS survey years. However, Amhara and Benishangul Gumuz regions showed an improvement in the ANCq from 2.35 and 2.64 in 2000 to 2.48 and 2.68 in 2016 EDHS years, respectively, and Addis Ababa and Dire Dawa cities have shown an AARC of 6.24 from the year 2000 to 2016 (**Figure 2**)

**Figure 2:**
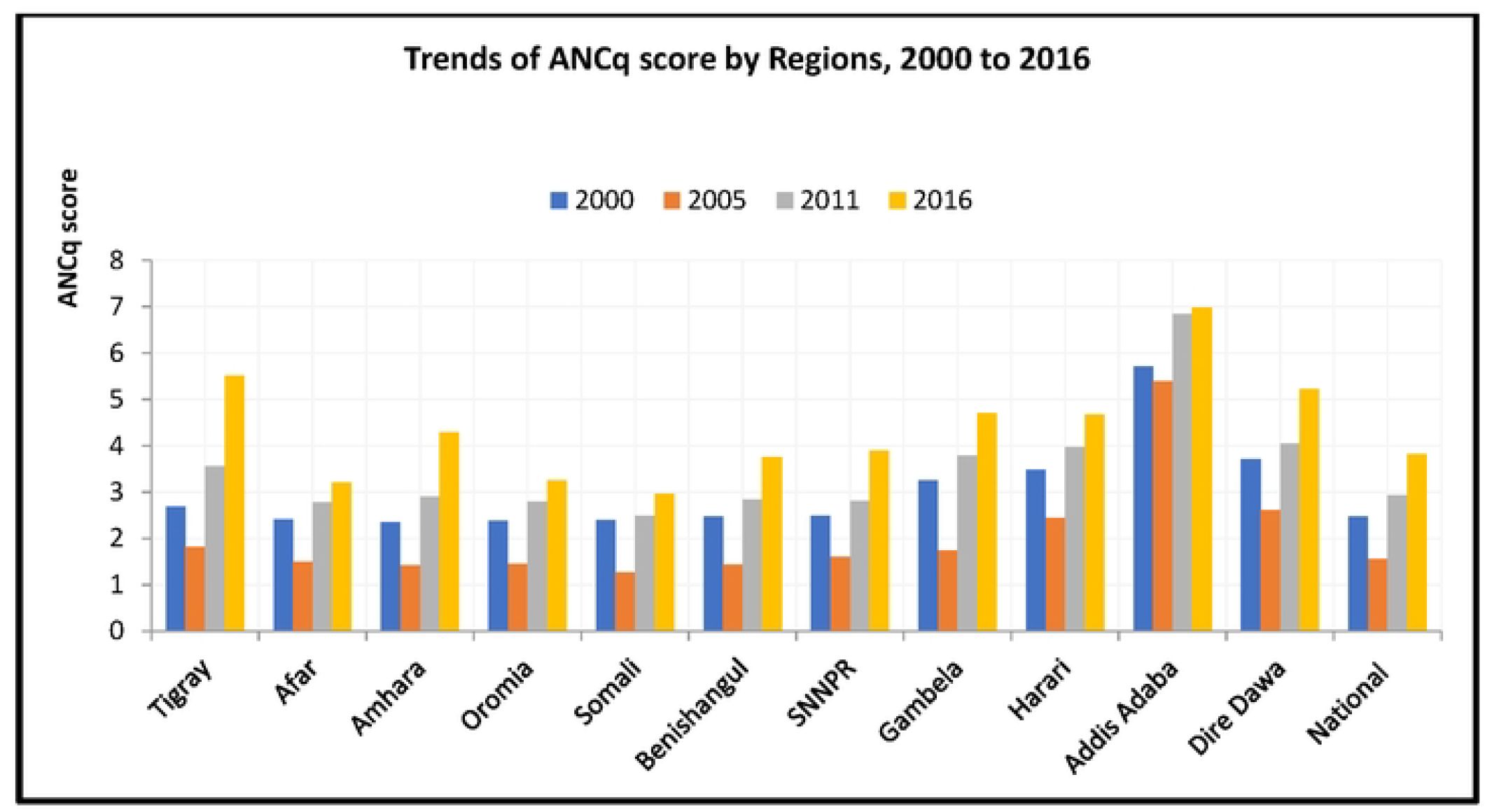
Trends of ANCq score across the regions from 2000-2016

Nationally, the ANCq score increased from 2.48 in 2000 to 2.69 in 2016, with an average annualized rate of increment (AARI) of 4.74%. Both Addis Ababa and Dire Dawa cities experienced average annual rates of increment of 1.55% and 1.83%, respectively, from 2000 to 2016. In contrast, the Amhara and Benishangul Gumuz regions saw improvements in their ANCq scores from 2.35 and 2.64 in 2000 to 2.48 and 2.68 in 2016, respectively. Notably, Addis Ababa and Dire Dawa cities recorded an AARC of 6.24 from 2000 to 2016 (**Figure 3**).

**Figure 3:**
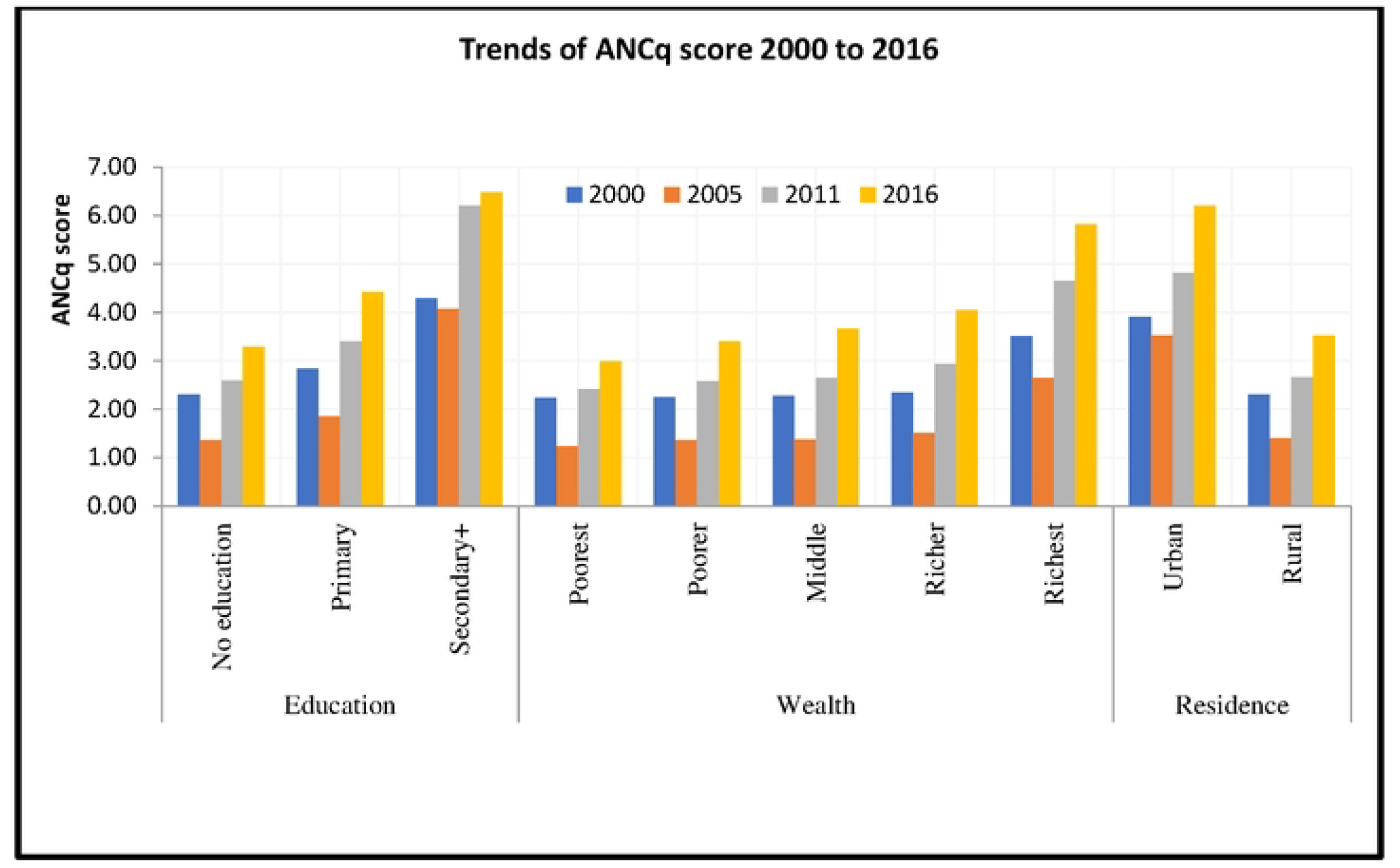
Trends of ANCq score across the different categories of residence, wealth, and education frorn 2000-2016

### Factors associated with neonatal mortality

**Table 3 describes** The association between maternal characteristics and neonatal mortality. Women who received four or more ANC visits (AN4+) had a 54% lower odds of neonatal mortality (AOR 0.46, 95% CI: 0.33, 0.64) compared to those with fewer than four visits. Additionally, women who received high-quality ANC had a 44% reduction in neonatal mortality (AOR 0.56, 95% CI: 0.42, 0.74) compared to those who received poor-quality care (**Table 3**).

**Table 3:**
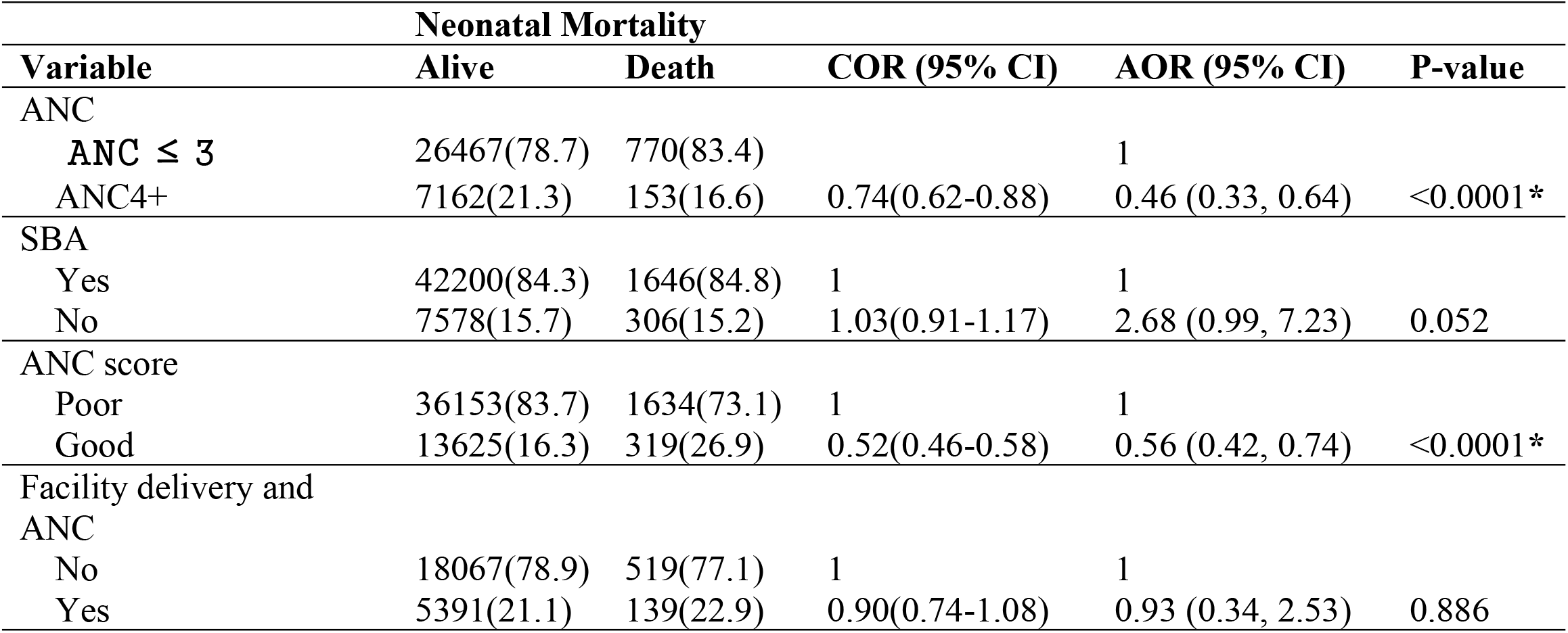
Factors associated with neonatal mortality among women attendants from 2000 to 2016 using multivariable logistic regression analysis.

**Table 4. Describes the association of ANC components with neonatal mortality**.

**Table 4:**
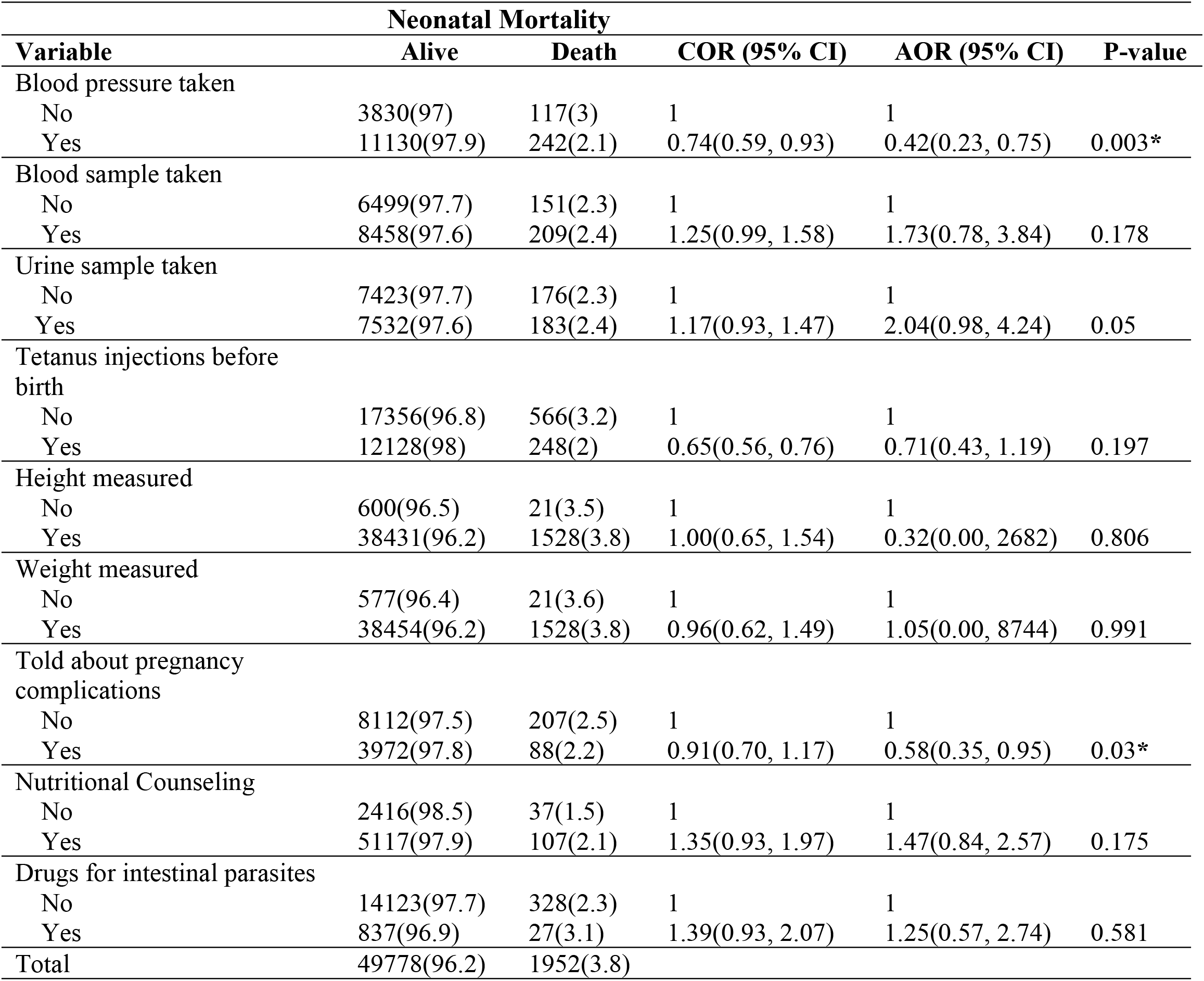
ANC component associated with neonatal mortality among women attendants from 2000 to 2016 using multivariable logistic regression analysis.

| Variable | Neonatal Mortality |  | COR (95% CI) | AOR (95% CI) | P-value |
| --- | --- | --- | --- | --- | --- |
|  | Alive | Death |  |  |  |
| Blood pressure taken |  |  |  |  |  |
| No | 3830(97) | 117(3) | 1 | 1 |  |
| Yes | 11130(97.9) | 242(2.1) | 0.74(0.59, 0.93) | 0.42(0.23, 0.75) | 0.003* |
| Blood sample taken |  |  |  |  |  |
| No | 6499(97.7) | 151(2.3) | 1 | 1 |  |
| Yes | 8458(97.6) | 209(2.4) | 1.25(0.99, 1.58) | 1.73(0.78, 3.84) | 0.178 |
| Urine sample taken |  |  |  |  |  |
| No | 7423(97.7) | 176(2.3) | 1 | 1 |  |
| Yes | 7532(97.6) | 183(2.4) | 1.17(0.93, 1.47) | 2.04(0.98, 4.24) | 0.05 |
| Tetanus injections before birth |  |  |  |  |  |
| No | 17356(96.8) | 566(3.2) | 1 | 1 |  |
| Yes | 12128(98) | 248(2) | 0.65(0.56, 0.76) | 0.71(0.43, 1.19) | 0.197 |
| Height measured |  |  |  |  |  |
| No | 600(96.5) | 21(3.5) | 1 | 1 |  |
| Yes | 38431(96.2) | 1528(3.8) | 1.00(0.65, 1.54) | 0.32(0.00, 2682) | 0.806 |
| Weight measured |  |  |  |  |  |
| No | 577(96.4) | 21(3.6) | 1 | 1 |  |
| Yes | 38454(96.2) | 1528(3.8) | 0.96(0.62, 1.49) | 1.05(0.00, 8744) | 0.991 |
| Told about pregnancy complications |  |  |  |  |  |
| No | 8112(97.5) | 207(2.5) | 1 | 1 |  |
| Yes | 3972(97.8) | 88(2.2) | 0.91(0.70, 1.17) | 0.58(0.35, 0.95) | 0.03* |
| Nutritional Counseling |  |  |  |  |  |
| No | 2416(98.5) | 37(1.5) | 1 | 1 |  |
| Yes | 5117(97.9) | 107(2.1) | 1.35(0.93, 1.97) | 1.47(0.84, 2.57) | 0.175 |
| Drugs for intestinal parasites |  |  |  |  |  |
| No | 14123(97.7) | 328(2.3) | 1 | 1 |  |
| Yes | 837(96.9) | 27(3.1) | 1.39(0.93, 2.07) | 1.25(0.57, 2.74) | 0.581 |
| Total | 49778(96.2) | 1952(3.8) |  |  |  |

Women who had blood pressure screening tests during ANC visits experienced a 58% reduction in neonatal mortality (AOR = 0.42, 95% CI: 0.23, 0.75) compared to those who did not receive the service. Similarly, women who were counseled and educated about pregnancy complications at any of their visits had a 42% lower risk of neonatal mortality (AOR = 0.58, 95% CI: 0.35, 0.95) compared to those who did not receive counseling services (**Table 4**).

## Discussion

This study examined the impact of antenatal care on neonatal mortality in Ethiopia using a nationally representative household survey conducted in 2000, 2005, 2011, 2016, and 2019. We calculated the components of basic routine antenatal care services and used the results to examine the antenatal care Quality Index. To understand how antenatal care coverage and ANC visits four and above affect neonatal mortality. The model for the demand for the quality of the antenatal care index is estimated using a logistic regression. Whereas the number of antenatal care visits, ANC quality index, and utilization of antenatal care services are estimated using logistic regression.

This study found that neonatal mortality significantly decreased from 58 to 36 deaths per 100,000 live births between 2000 and 2019. Women with four or more ANC visits were 54% less likely to experience neonatal mortality compared to those with fewer visits. Frequent and timely ANC follow-up is essential in obstetric care, particularly in low- and middle-income countries like Ethiopia. This finding was consistent with the studies conducted in different parts of Ethiopia (16,17).

The findings of the study were similar to the study carried out in Zimbabwe, where it was found that having ANC follow-up had a significant (negative) effect on the likelihood of neonatal mortality (15). The results are also consistent with a study done in Sub-Saharan Africa, which found that women who receive antenatal care follow-up have a lower risk of neonatal mortality (5). Similarly, studies conducted in Indonesia (16), India, Bangladesh (18), and Brazil (19) also reported that ANC has a significant effect on the reduction of neonatal mortality.

A previously conducted meta-analysis reported a strong negative association between perinatal mortality and lack of antenatal care (18). This is because women who receive ANC services, including pregnancy checkups, health promotion, and disease prevention like iron/folic acid supplementation, tetanus vaccination, and counseling by skilled providers, significantly improve neonatal survival. Frequent ANC visits increase access to quality obstetric care, thereby reducing the likelihood of neonatal mortality. This is important to detect any deviation from normality so that problems are detected early, and appropriate measures can be taken promptly. In addition, ANC has indirect benefits since women attending ANC are more likely to have their delivery assisted by a skilled birth attendant or give birth in health facilities (15).

Similarly, the odds of receiving a good quality of antenatal care were associated with a lower risk of neonatal mortality by nearly 44% as compared to a poor level of antenatal care quality. This finding was supported by a study conducted in Zimbabwe, which revealed that a one-unit increase in the quality of antenatal care decreased the risk of neonatal mortality by nearly 42.3% (8). Similarly, a result from pooled estimates of meta-analysis from 60 low- and middle-income countries showed that the risk of neonatal mortality is lower with the ANC quality index increase. It is well-known that ANC visits may help to reinforce maternal education and compliance and provide an opportunity for screening for warning signs of pregnancy complications and treatment of infections (19).

In addition, ANC provides an important opportunity for health workers to teach mothers how to recognize warning signs of complications during pregnancy, labor, and delivery, and encourage them to plan clean and safe deliveries, preferably with trained assistance (20). Another reason could be that if the quality of care improves, more women will likely attend prenatal care appointments and have skillful deliveries, which could reduce the neonatal mortality rate.

This comprehensive analysis also tried to look at the effect of individual antenatal care components on the risk of neonatal mortality. According to the pooled data for the years 2000 to 2016, women having blood pressure screening tests during ANC visits lowers the risk of neonatal mortality by 58% for women who didn’t get the service. This finding is congruent with a study conducted in Zimbabwe, which indicated that mothers who receive blood pressure checks during pregnancy are less likely to experience neonatal mortality (8).

Routine blood pressure checks are an essential component of antenatal care visits since high blood pressure during pregnancy poses numerous risks to the mother and the unborn child. For example, high blood pressure during pregnancy might result in decreased blood flow to the placenta, which restrains the movement of oxygen to the baby and potentially retards growth. High blood pressure might also result in premature delivery and low birth weight babies, which all contribute to adverse health outcomes, including infant death (19).

Similarly, women who had been counseled and educated about pregnancy complications at any of their visits were statistically associated with a lower risk of neonatal mortality compared with women who didn’t receive counseling services. The possible justification would be that women who had received counseling services during ANC follow-up would apply prevention mechanisms, including facility delivery, and post-natal care follow-up. The explanation could be that women who received counseling services about pregnancy complications during ANC follow-up would use preventative measures, such as facility birth and post-natal care follow-up, which would have a great contribution to decreasing neonatal mortality.

We acknowledge that the study is confined to each woman’s most recent birth, which happened during the five years before the survey for which we observe antenatal care data. This data limitation could be problematic because it makes it harder to generalize the findings to a broader context. It would be interesting to investigate the effects of within-women disparities in antenatal care utilization on siblings’ survival probability.

Furthermore, the restriction to the most recent birth could result in sample selection bias, which could potentially affect our estimates. However, since these limitations are a result of the data collection methodology adopted by MEASURE DHS, we are unable to examine any within-sibling differences in survival. Lastly, we ought to emphasize that our estimates might still be minimally biased by potential endogeneity resulting from the voluntary nature of the antenatal care decision. Besides the noted concerns, our analysis adds a significant contribution to the recent discussions in less-developed countries on the importance of antenatal care for child health outcomes.

The result of this study showed that the quality of antenatal care reduces the risk of neonatal mortality. This implies that a substantial improvement in the quality of ANC services is necessary to improve the survival of neonates. As the ANC contact increases 4 times or more through the pregnancy period, it reduces the likelihood of neonatal mortality. To reduce neonatal deaths and achieve the SDG30 goal, all pregnant women should receive comprehensive antenatal care. Educating mothers on the importance of antenatal visits and ensuring the management of maternal conditions can reduce neonatal mortality. Public health decision-makers must ensure pregnant women receive essential antenatal care components.

Moreover, during ANC visits, strong emphasis is required to ensure that the health care provider provides counseling on complications related to pregnancy, to delivery in the health facility, which may reduce neonatal mortality by preventing sepsis and managing and treating preterm birth.

## Data Availability

All relevant data are within the manuscript and its Supporting Information files

https://dhsprogram.com/data/dataset/Ethiopia_Standard-DHS_2016.cfm

https://dhsprogram.com/data/dataset/Ethiopia_Standard-DHS_2011.cfm

https://dhsprogram.com/data/dataset/Ethiopia_Standard-DHS_2005.cfm

https://dhsprogram.com/data/dataset/Ethiopia_Standard-DHS_2000.cfm

## Abbreviations

AARC: Average Annualized Rate of Change
ANC: Antenatal Care
AOR: Adjusted Odds Ratio
CI: Confidence Intervals
COR: Crude Odds Ratio
EDHS: Ethiopian Demographic and Health Surveys

## Acknowledgments

We gratefully acknowledge the financial support provided by Countdown to 2030 for the preparation of this manuscript.

## Data availability

All data are fully available and included within the manuscript and Supporting Information files.

## Author Contributions

**Conceptualization:** GF, TA, GM

**Data curation:** GF, TA, GM, AA

**Formal analysis:** GF, TA, GM, AA

**Methodology:** GF, TA, GM, AA

**Supervision:** GF, TA, GM, AA, AH

**Validation:** GF, TA, GM,AA, AH,MS

**Visualization:** GF, TA, GM, AA, AH, MS

**Writing** – original draft: GF, TA, GM, AA, AH

Writing – review & editing: All

## Ethics approval

Ethical approval was obtained from the Ethiopian Public Health Institute Institutional Review Board.

## Author Details

^1^ Ethiopian Public Health Institute (EPHI), Addis Ababa, Ethiopia.

## References

1. Alebel A, Wagnew F, Petrucka P, Tesema C, Moges NA, Ketema DB, et al. Neonatal mortality in the neonatal intensive care unit of Debre Markos referral hospital, Northwest Ethiopia: A prospective cohort study. BMC Pediatr. 2020;20(1):1–11.

2. World Health Organization. World health statistics 2024. Geneva: WHO; 2024. Report No.: ISBN 9789240094703. p. 1–96.

3. UNICEF. Levels and trends in child mortality 2019. New York: United Nations Children’s Fund; 2019. p. 1–52.

4. Ethiopian Public Health Institute (EPHI), ICF. Ethiopia mini demographic and health survey 2019: Final report. Addis Ababa, Ethiopia, and Rockville, Maryland, USA: EPHI and ICF; 2021. p. 1–207. Available from: https://dhsprogram.com/pubs/pdf/FR363/FR363.pdf

5. Wondemagegn AT, Alebel A, Tesema CT, Ketema WA. The effect of antenatal care on perinatal outcomes in Ethiopia: A systematic review and meta-analysis. PLoS One. 2021;16(1):e0244878.

6. Singh A, Pallikadavath S, Ram F, Alagarajan M. Do antenatal care interventions improve neonatal survival in India? Health Policy Plan. 2014;29(7):842–8.

7. Garrido G. The impact of adequate prenatal care in a developing country: Testing the WHO recommendations. [dissertation]. 2009.

8. Makate M, Makate C. The impact of prenatal care quality on neonatal, infant, and child mortality in Zimbabwe: Evidence from the demographic and health surveys. Health Policy Plan. 2017;32(3):395–404.

9. World Health Organization. World health statistics 2015. Geneva: WHO; 2015.

10. United Nations Inter-agency Group for Child Mortality Estimation (UN IGME). Levels and trends in child mortality 2021. New York: United Nations Children’s Fund; 2021. p. 1–70. Available from: https://data.unicef.org/resources/levels-and-trends-in-child-mortality-2021/

11. Lambon-Quayefio MP, Owoo NS. Examining the influence of antenatal care visits and skilled delivery on neonatal deaths in Ghana. Appl Health Econ Health Policy. 2014;12(5):511–22.

12. World Health Organization. WHO recommendations on antenatal care for a positive pregnancy experience. Geneva: WHO; 2016.

13. The DHS Program. Demographic and Health Survey (DHS). 2025. Available from: https://dhsprogram.com/methodology/survey-Types/dHs.cfm

14. Croft TN, Marshall AMJ, Allen CK. Guide to DHS statistics. Rockville, Maryland, USA: ICF; 2018.1–90.

15. Deb P, Sosa-Rubi SG. Does onset or quality of prenatal care matter more for infant health? New York: City University of New York 2005.

16. Titaley CR, Dibley MJ. Antenatal iron/folic acid supplements, but not postnatal care, prevent neonatal deaths in Indonesia: Analysis of Indonesia Demographic and Health Surveys 2002/2003–2007. BMJ Open. 2012;2(6):e001399.

17. Mersha A, Bante A, Shibiru S. Neonatal mortality and its determinants in public hospitals of Gamo and Gofa zones, southern Ethiopia: Prospective follow-up study. BMC Pediatr. 2019;19(1):1–8.

18. Islam MA, Biswas B. Socio-economic factors associated with increased neonatal mortality: A mixed-method study of Bangladesh and 20 other developing countries based on demographic and health survey data. Clin Epidemiol Glob Health. 2021;11:100801.

19. Kale PL, De Mello-Jorge MHP, Da Silva KS, Fonseca SC. Near miss and neonatal death cases: Factors associated with life-threatening conditions in six maternity hospitals in Southeast Brazil. Cad Saude Publica. 2017;33(4):e00105316.

20. Bloom SS, Lippeveld T, Wypij D. Does antenatal care make a difference to safe delivery? A study in urban Uttar Pradesh, India. Health Policy Plan. 1999;14(1):38–48.

